# Intraprocedural Doppler and Invasive Hemodynamic Profiling Predict Clinical Outcomes After Mitral TEER

**DOI:** 10.1101/2023.04.01.23288045

**Authors:** Syed Zaid, Priscilla Wessly, Taha Hatab, Safi U Khan, Nadeen Faza, Stephen H Little, Marvin D Atkins, Michael J Reardon, Neal S Kleiman, William A Zoghbi, Sachin S Goel

## Abstract

**Background:** Whether intraprocedural changes in left atrial pressure and Doppler Echocardiographic parameters are synergistic in predicting outcomes after mitral transcatheter edge-to-edge repair (TEER) is not currently known. We sought to evaluate real-time changes in invasive hemodynamics and non-invasive Doppler to develop intraprocedural profiles and assess their impact on clinical outcomes after TEER for MR.

**Methods:** Intraprocedural changes in hemodynamics and Doppler flow with transesophageal echocardiography were assessed in 181 patients with significant MR (51.9% primary MR) undergoing TEER between 2014 and 2022. Independent predictors of the primary composite endpoint of 1-year mortality and heart failure hospitalization (HFH) were identified using multivariable Cox-regression. With receiver operating characteristic curve-derived thresholds for the predictors of the primary end-point, patients were stratified into hemodynamic profiles based on the number of predictors present, and their impact on outcomes was examined.

**Results:** Median follow-up was 21.3 months (IQR:11.3-36.5), with 1-year mortality and HFH rates of 19.3% and 12.7%, respectively. Residual mean left atrial pressure (mLAP) [HR=1.073/mmHg (1.03-1.12)], a lesser degree of MR reduction [HR=0.65/grade (0.45-0.93)], and lesser increment in PV systolic time velocity integral (S-VTI) [HR=0.95/cm (0.91-0.99)] were independent predictors of 1-year mortality/HFH. MR reduction by <3 grades (33.1%), S-VTI increment ≤8cm (33.9%), and residual mLAP >15mmHg (43.6%) were the most predictive thresholds. Optimal profile (0 predictors), Mixed (1 predictor) and Poor profile (≥2 predictors) were present in 28.7%, 39.2% and 32.0% of cases respectively. Two-year cumulative event-free survival was 60.1% overall, and higher in patients with optimal profile compared to mixed/poor groups (84.7% vs 55.5% vs 43.3%, P<0.001). There was an incremental risk of mortality/HFH with each profile overall [HR=1.75/profile (1.34-2.29)], and within primary MR [HR=1.64/profile (1.15-2.36)] and secondary MR [HR=1.77/profile (1.17-2.68)] cohorts. There was also an incremental risk of mortality alone with each profile [HR=1.65/profile (1.22-2.22)]. Hemodynamic profile was an independent predictor of 1-year mortality [HR=1.98/profile (1.21-3.25)] after TEER, along with baseline tricuspid regurgitation severity [HR=1.55/grade (1.10-2.19)], and post-procedural transmitral mean gradient>5mmHg [HR=2.32 (1.17-4.61)].

**Conclusion:** Intraprocedural hemodynamic profiling integrating changes in invasive hemodynamics and non-invasive doppler provide prognostic information in patients undergoing TEER and may provide real-time intraprocedural guidance to optimize long-term clinical outcomes.

## Introduction

Mitral transcatheter edge-to-edge repair (TEER) has proven to be a beneficial therapy for high surgical risk patients with primary mitral regurgitation (MR) and for patients with secondary MR who remain symptomatic with significant MR despite optimization of guideline directed medical therapy (GDMT).^1, 2^ With more than 150,000 mitral TEER performed worldwide, identifying patients that benefit most from this therapy has been a matter of ongoing research. Favorable outcomes after mitral TEER have been associated with mild residual MR, primary MR with no left ventricular (LV) dilatation or right ventricular (RV) dysfunction, and lower baseline serum creatinine among others.^3–5^ Conversely, the most consistently reported predictors of worse outcomes include secondary MR, atrial fibrillation, low leaflet-to-annulus index, large left atrial volume index, severe tricuspid regurgitation, and pulmonary hypertension.^5–9^ In a recent study that stratified patients undergoing TEER based on residual MR and mean left atrial pressure (LAP), 1-year survival was superior among patients with optimal reduction in MR and normal postprocedural LAP.^10^ Intraprocedural transesophageal echocardiography (TEE) derived pulmonary vein (PV) Doppler is emerging as a useful adjunct to assess MR reduction during the procedure, and few studies thus far have investigated the impact of changes in PV Doppler on outcomes after mitral TEER.^11, 12^ However, no prior studies have integrated real-time changes in non-invasive doppler such as TEE-derived PV flow and invasive hemodynamics to study long term outcomes after TEER. We therefore sought to investigate the clinical significance of changes in invasive hemodynamics and Doppler parameters with TEER, and test whether we could develop intraprocedural multiparametric profiles that have prognostic impact after Mitral TEER.

## Patients and Methods

### Data Source

We retrospectively reviewed 305 consecutive patients with moderate-severe or severe MR who underwent mitral TEER with MitraClip (Abbott Vascular, Santa Clara, CA) at Houston Methodist Hospital (Houston, Texas, USA) from March 2014 to March 2022. As determined by a multidisciplinary heart team based on current guidelines, patients with symptomatic primary MR at high surgical risk and those with secondary MR on optimized GDMT respectively underwent the procedure. Among 286 patients (94%) who had successful MitraClip implantation, 181 patients had hemodynamics, analyzable PV flow Doppler and color Doppler MR evaluation on intraprocedural TEE before and after the procedure and comprised the primary study cohort. All study procedures were conducted in accordance with the Declaration of Helsinki. This observational study was approved by the Houston Methodist Institutional Review Board.

### Invasive Hemodynamics

The procedure was performed under general anesthesia with TEE and fluoroscopic guidance. After a transseptal puncture, a 24-F transseptal sheath was used to measure LAP and v-wave at baseline prior to clip delivery system insertion. LAP and v-wave were continuously monitored during the procedure. After the final clip deployment, direct LAP and v-wave were measured before withdrawal of the sheath from the LA to the right atrium.

### Echocardiographic Analysis

All patients had pre-procedural TTE and TEE using Philips’s echocardiography system (i33 instruments; Philips Technology, Amsterdam, The Netherlands). American Society of Echocardiography guidelines were used to assess the mechanism and severity of MR as mild (1+), moderate (2+), moderate to severe (3+), and severe (4+).^13^ The mechanism of MR was classified as primary/degenerative, secondary, or mixed based on guidelines.^13^

Intraprocedural TEE was performed to confirm the severity of MR and obtain mean mitral gradient using continuous wave Doppler of the mitral inflow tracing. In addition, PV flow was obtained from the right and left upper pulmonary veins by pulse wave Doppler. The sample volume was placed 1 cm from the ostium of the right and left upper pulmonary vein. TEE images were reviewed, and velocity time integrals (VTI) and peak velocities of the systolic wave (S-VTI, Sv) and the diastolic wave (D-VTI, Dv) were determined, in addition to the MR grade and mean transmitral gradient. The PV with the lowest or most negative Sv was chosen.^14^ The same vein was used to assess the change in flow after MitraClip implantation. PV flow patterns were classified as reversed, normal and blunted as described previously.^12^ Finally, the change in each of the four PV Doppler variables (S-VTI, D-VTI, Sv, Dv) with MitraClip was determined and defined as ΔS-VTI, ΔD-VTI, ΔSv and ΔDv respectively **(Figure 1).**

**Figure 1.**
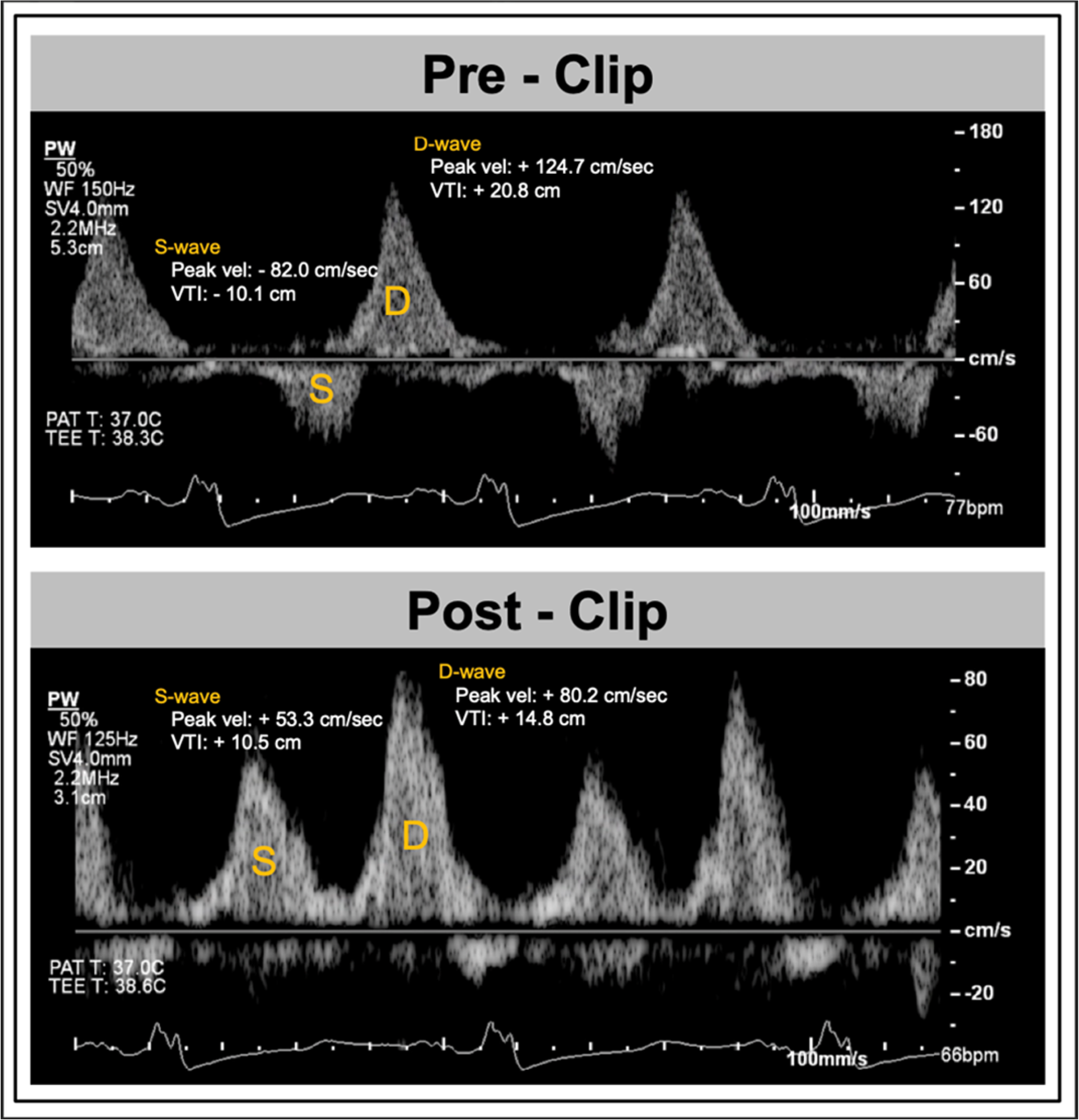
Assessment of Pulmonary Venous Flow using intraprocedural TEE. Pulmonary venous (PV) flow before and after successful MitraClip implantation using transesophageal echo derived pulsed wave doppler of the left upper PV. PV flow shows systolic flow reversal at baseline, with upright systolic wave after MitraClip implantation associated with an increase in S-VTI of 20 cm.

### Statistical Analysis

Clinical, procedural, and echocardiographic characteristics were collected for all patients before and after the mitral TEER procedure. Depending on the distribution of data, continuous variables are reported as means with standard deviation or median with IQR, whereas categorical variables are reported as frequencies and proportions. The Kolmogorov-Smirnov test was used for assessment of normality for continuous data. The primary endpoint was composite of all-cause mortality and heart failure hospitalization (HFH) at 1-year. Univariate Cox regression analysis was used to identify variables associated with 1-year mortality/HFH. Since model building was limited by the relative number of mortality events, forward, stepwise, multivariable Cox regression models were developed. All variables with p<0.10 from univariate analysis in addition to the most predictive PV variable influencing the outcomes of interest were considered for the multivariable Cox regression analysis, and only those with p<0.05 were included in the final model. Using receiver operating characteristic (ROC) curve-derived thresholds for the independent predictors of the primary end-point, patients were stratified into hemodynamic profiles based on number of predictors present.

Hemodynamic profiles were defined as optimal (no predictors), mixed (1 predictor), and poor (≥2 predictors) **(Figure 2)**. Differences between the three profiles were detected using the analysis of variance for continuous variables, and chi-squared or Fisher’s exact test for categorical variables. Profile-specific changes in intraprocedural hemodynamic and TEE-derived PV variables with MitraClip were assessed using paired-*t* test, and analysis of covariance was performed to detect differences in mean change between the three profiles, while adjusting for the baseline values. Kaplan-Meier analysis was used to assess survival estimates for the primary endpoint in the overall population and stratified by Primary and Secondary MR, and differences in survival between profiles were compared using the log-rank test. Impact of hemodynamic profile on the endpoints was determined using Cox regression analysis. A two-sided *p<* 0.05 was considered statistically significant and all statistical analyses were performed using SPSS version 24.0 (IBM, Armonk, New York).

**Figure 2.**
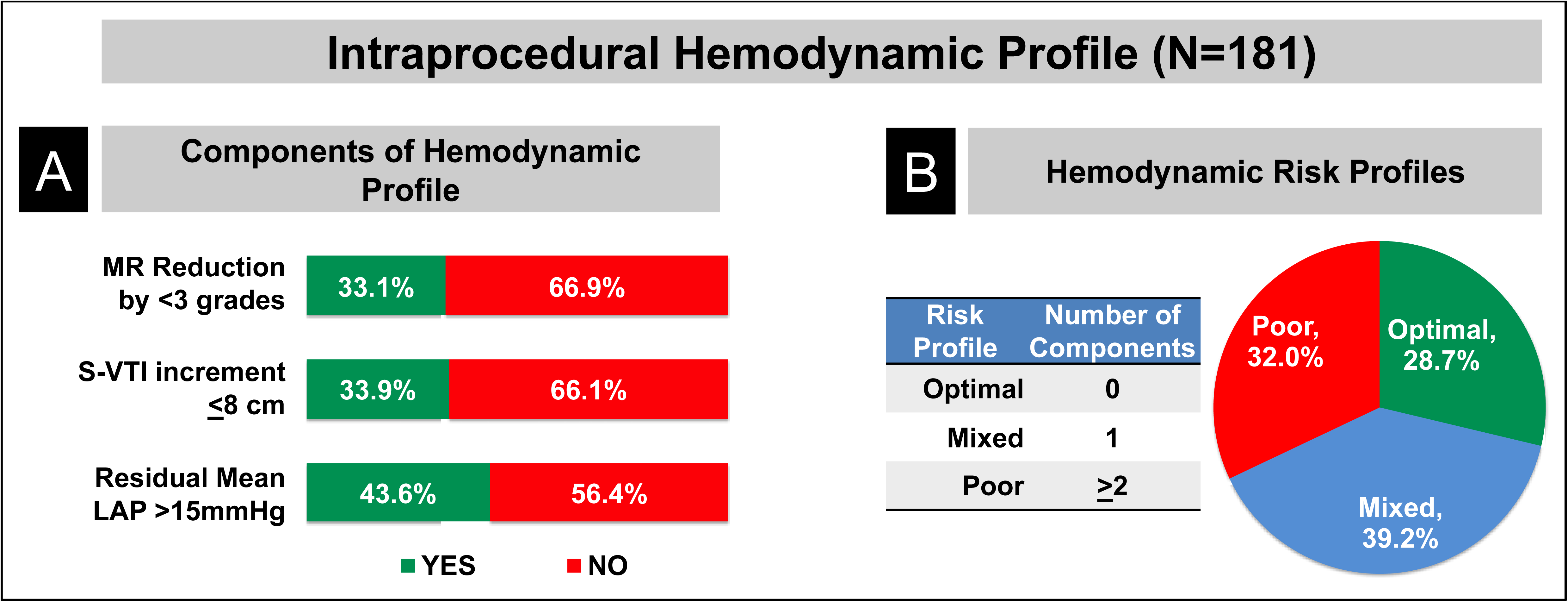
Intraprocedural Hemodynamic Profile. Mitral Regurgitation by <3 Grades, Change in Systolic Velocity-Time-Integral by ≤8 cm and residual mean Left Atrial Pressure of >15 mmHg were the three components of the Intraprocedural Hemodynamic Profile (A). Hemodynamic profiles were defined as optimal, mixed, or poor if 0, 1 or ≥2 of these components were present (B).

## Results

### Baseline Clinical, Echocardiographic and Procedural Characteristics

Of the 181 patients (mean age 76.4±10.6 years, 42.5% female) included in our final analysis, 94 (51.9%) patients had primary MR, 69 (38%) patients had secondary MR while 18 (10%) patients had MR of mixed etiology. All patients had MR grade ≥3+ with the vast majority having 4+ (83%) MR. NYHA III/IV was present in 138 (76.7%) patients. Baseline clinical and echocardiographic characteristics are listed in **Table 1**.

**TABLE 1.** Baseline Clinical and Echocardiographic Characteristics.

|  | **Overall**<br>**N=181** | **Optimal**<br>**N=52** | **Mixed**<br>**N=71** | **Poor**<br>**N=58** | *p value* |
| --- | --- | --- | --- | --- | --- |
| **Clinical Characteristics** |  |  |  |  |  |
| Age (years) | 76.4 [10.6] | 79.3 [10.0] | 77.1 [8.7] | 73.1 [12.3] | **0.006** |
| Female | 77 (42.5) | 23 (44.2) | 28 (39.4) | 26 (44.8) | 0.79 |
| Hypertension | 136 (75.1) | 41 (78.8) | 52 (73.2) | 43 (74.1) | 0.76 |
| Atrial Fibrillation | 104 (57.8) | 30 (57.7) | 43 (60.6) | 31 (54.4) | 0.78 |
| Coronary artery disease | 70 (38.7) | 20 (38.5) | 27 (38) | 23 (39.7) | 0.98 |
| Frailty | 116 (64.1) | 28 (53.8) | 49 (69) | 39 (67.2) | 0.19 |
| Diabetes | 54 (29.8) | 13 (25) | 18 (25.4) | 23 (39.7) | 0.14 |
| Prior Stroke | 20 (11) | 3 (5.8) | 7 (9.9) | 10 (17.2) | 0.15 |
| Dialysis | 15 (8.3) | 2 (3.8) | 5 (7) | 8 (13.8) | 0.15 |
| Prior MI | 35 (19.3) | 16 (30.8) | 9 (12.7) | 10 (17.2) | **0.038** |
| Prior Pacemaker | 25 (13.8) | 9 (17.3) | 11 (15.5) | 5 (8.6) | 0.37 |
| Prior ICD | 30 (16.6) | 6 (11.5) | 11 (15.5) | 13 (22.4) | 0.30 |
| Prior CABG | 44 (24.3) | 16 (30.8) | 16 (22.5) | 12 (20.7) | 0.42 |
| Prior PCI | 44 (24.3) | 11 (21.2) | 17 (23.9) | 16 (27.6) | 0.73 |
| BMI (kg/m<sup>2</sup>) | 26.1 [6] | 25.4 [4.6] | 25.6 [4.9] | 27.2 [8] | 0.20 |
| BSA (m<sup>2</sup>) | 1.9 [0.3] | 1.9 [0.2] | 1.9 [0.3] | 1.9 [0.3] | 0.80 |
| NYHA Class |  |  |  |  | 0.066 |
| I | 7 (3.9) | 5 (9.8) | 2 (2.8) | 0 (0) |  |
| II | 35 (19.4) | 9 (17.6) | 10 (14.1) | 16 (27.6) |  |
| III | 122 (67.8) | 32 (62.7) | 54 (76.1) | 36 (62.1) |  |
| IV | 16 (8.9) | 5 (9.8) | 5 (7.0) | 6 (10.3) |  |
| KCCQ12 score | 43.7 [26.9] | 47.5 [25.1] | 44.4 [29.3] | 39.6 [25] | 0.36 |
| STS Risk MV replacement (%) | 7.2 [5.4] | 7.3 [5.2] | 6.7 [5] | 7.7 [6.1] | 0.60 |
| STS Risk MV repair (%) | 5.2 [4.8] | 5.7 [4.2] | 4.9 [4.6] | 5.3 [5.6] | 0.68 |
| **Echocardiographic Characteristics** |  |  |  |  |  |
| MR Etiology |  |  |  |  | 0.07 |
| Primary | 90 (50.7) | 33 (66) | 37 (53.5) | 20 (34.4) |  |
| Secondary | 58 (32.8) | 11 (22) | 21 (30.4) | 26 (44.8) |  |
| Atrial FMR | 11 (6.2) | 2 (4) | 5 (7.2) | 4 (6.9) |  |
| Mixed | 18 (10.2) | 4 (8) | 6 (8.7) | 8 (13.8) |  |
| MR Severity |  |  |  |  | **<0.001** |
| Moderate-Severe | 31 (17.1) | 2 (3.8) | 8 (11.3) | 21 (36.2) |  |
| Severe | 150 (82.9) | 50 (96.2) | 63 (88.7) | 37 (63.8) |  |
| MAC Severity |  |  |  |  | 0.24 |
| None | 124 (68.5) | 35 (67.3) | 48 (67.6) | 41 (70.7) |  |
| Mild | 32 (17.7) | 13 (25) | 12 (16.9) | 7 (12.1) |  |
| Moderate | 15 (8.3) | 2 (3.8) | 5 (7) | 8 (13.8) |  |
| Severe | 10 (5.5) | 2 (3.8) | 6 (8.5) | 2 (3.4) |  |
| TR Severity |  |  |  |  | 0.20 |
| None/Trace | 44 (24.3) | 14 (26.9) | 18 (25.4) | 12 (20.7) |  |
| Mild | 65 (35.9) | 21 (40.4) | 30 (42.3) | 14 (24.1) |  |
| Moderate | 58 (32) | 14 (26.9) | 19 (26.8) | 25 (43.1) |  |
| Severe | 14 (7.7) | 3 (5.8) | 4 (5.6) | 7 (12.1) |  |
| PASP (mmHg) | 47.7 [17.9] | 48.4 [18.7] | 47 [19.3] | 47.7 [16] | 0.97 |
| LVEF (%) | 51.2 [14.8] | 55.7 [13.2] | 51.7 [15.3] | 46.6 [14.4] | **0.005** |
| LVIDs (cm) | 3.9 [1.2] | 3.7 [1.2] | 3.7 [1.2] | 4.3 [1.1] | **0.009** |
| LVIDd (cm) | 5.4 [1] | 5.3 [0.9] | 5.2 [1] | 5.5 [1] | 0.21 |
| LA Volume (mL) | 122.6 [58.2] | 115.3 [52.5] | 126.7 [55.4] | 123.6 [66.2] | 0.57 |
| LAVI (mL/m<sup>2</sup>) | 65.1 [28.1] | 61.3 [24.4] | 68.1 [28.1] | 64.8 [31.1] | 0.44 |

Values are expressed as Mean [Standard Deviation] or N (%)
BMI = body mass index; BSA = body surface area; CABG = coronary artery bypass graft; ICD = Implantable cardioverter/defibrillator; KCCQ-12 = Kansas City Cardiomyopathy Questionnaire score; LAVI = left atrium volume index; LVEF = left ventricle ejection fraction; LVIDs/d = left ventricle internal diameter (systole/diastole); MAC = mitral annular calcification; MI = myocardial infarction; MV = mitral valve; NYHA = New York Heart Association; PASP = pulmonary artery systolic pressure; PCI = percutaneous coronary intervention; RAP = right atrial pressure, STS = Society for Thoracic Surgeons.

### Procedural Characteristics

Procedural details are summarized in **Table 2**. Total number of clips deployed averaged 1.5±0.6 per patient, with 54.7% receiving 1 MitraClip, and 45.3% patients receiving >1 device. MR reduction to ≤2+ MR was achieved in 97.8% at the end of the procedure, with a mean reduction of 2.8±0.7 grades. At baseline, PV systolic reversal was the predominant morphology (59.7%) with S-dominance seen in only 1 patient. However, after MitraClip implantation, no patients had PV systolic reversal with S-dominance seen in 55.6% of cases. Overall, mitral TEER was associated with a significant increment in TEE-derived S-VTI and peak Sv, and reduction in D-VTI and peak Dv (all p<0.001), in addition to a significant reduction in invasive mLAP (from 20.0±7.8 mmHg to 14.9±5.7 mmHg, p<0.001) and v-wave (from 35.6±17.4 cm/s to 22.0±9.4 cm/s, p<0.001) (**Supplementary Table S1**).

**Table 2.** Procedural Characteristics.

|  | **Overall**<br>**N=181** | **Optimal**<br>**N=52** | **Mixed**<br>**N=71** | **Poor**<br>**N=58** | *p value* |
| --- | --- | --- | --- | --- | --- |
| Number of clips | 1.5 [0.6] | 1.4 [0.5] | 1.5 [0.6] | 1.6 [0.7] | 0.38 |
| 1 Clip | 99 (54.7) | 31 (59.6) | 37 (52.1) | 31 (53.4) | 0.27 |
| 2 Clips | 74 (40.9) | 21 (40.4) | 31 (43.7) | 22 (37.9) |  |
| >2 Clips | 8 (4.4) | 0 (0) | 3 (4.2) | 5 (8.6) |  |
| Type of MitraClip |  |  |  |  |  |
| Original MitraClip | 52 (28.7) | 17 (32.7) | 22 (31.0) | 13 (22.4) | 0.43 |
| NT | 12 (6.6) | 4 (7.7) | 3 (4.2) | 5 (8.6) | 0.57 |
| XT | 11 (6.1) | 3 (5.8) | 3 (4.2) | 5 (8.6) | 0.58 |
| NTR | 21 (11.6) | 7 (13.5) | 9 (12.7) | 5 (8.6) | 0.69 |
| XTR | 40 (22.1) | 10 (19.2) | 18 (25.4) | 12 (20.7) | 0.69 |
| NTW | 32 (17.7) | 9 (17.3) | 13 (18.3) | 10 (17.2) | 0.98 |
| XTW | 46 (25.4) | 13 (25.0) | 14 (19.7) | 19 (32.8) | 0.24 |
| Fluoroscopy time (mins) | 24.0 [14.0] | 24.4 [14.1] | 23.2 [12.8] | 24.4 [15.4] | 0.87 |
| PV wave morphology |  |  |  |  |  |
| Pre-Clip |  |  |  |  |  |
| S dominant | 1 (0.6) | 0 (0) | 1 (1.4) | 0 (0) | <0.001 |
| S blunting | 72 (39.8) | 6 (11.5) | 26 (36.6) | 40 (69) |  |
| S reversal | 108 (59.7) | 46 (88.5) | 44 (62) | 18 (31) |  |
| Post-Clip |  |  |  |  |  |
| S dominant | 100 (55.6) | 32 (61.5) | 42 (59.2) | 26 (45.6) | 0.27 |
| S blunting | 80 (44.5) | 20 (38.5) | 29 (40.8) | 31 (54.4) |  |
| S reversal | 0 (0) | 0 (0) | 0 (0) | 0 (0) |  |
| MR Severity (post-clip) |  |  |  |  |  |
| None/trace | 32 (17.7) | 14 (26.9) | 12 (16.9) | 6 (10.3) | <0.001 |
| Mild | 112 (61.9) | 38 (73.1) | 47 (66.2) | 27 (46.6) |  |
| Moderate | 33 (18.3) | 0 (0) | 11 (15.5) | 22 (37.9) |  |
| Moderate-severe | 4 (2.2) | 0 (0) | 1 (1.4) | 3 (5.2) |  |
| Severe | 0 (0) | 0 (0) | 0 (0) | 0 (0) |  |
| MR reduction (grade) | - 2.8 [0.7] | - 3.2 [0.4] | - 2.9 [0.6] | - 2.2 [0.8] | 0.21* |
| Hemodynamic Profile components |  |  |  |  |  |
| MR reduction by<br><3 grades | 60 (33.1) | 0 (0) | 18 (25.4) | 42 (72.4) | **<0.001** |
| S-VTI increment<br>≤ 8 cm | 61 (33.7) | 0 (0) | 19 (26.8) | 42 (72.4) | **<0.001** |
| Residual mLAP<br>>15 mmHg | 79 (43.6) | 0 (0) | 34 (47.9) | 45 (77.6) | **<0.001** |

Values are expressed as Mean [Standard Deviation] or N (%)
\* P calculated using Analysis of Covariance to adjust for pre-clip values.
D-VTI= diastolic velocity time integral; LAP= left atrial pressure; S-VTI= systolic velocity time integral; Δ= Difference between pre- and post-clip deployment

### Outcomes

Outcomes after Mitral TEER are summarized in **Table 3**. There was no in-hospital mortality, and median length of stay was 1 day (IQR: 1-2). MR grade ≤2+ was observed in 93.4% at 30-days, and 95% at 1-year follow up (**SupplementaryFigure S1**). At 30-days, there were 4 (2.2%) deaths and 5 (2.8%) HFH. Four patients (2.2%) had MV re-intervention within the first year, of which 2 had redo-MitraClip and 2 underwent MV surgery. Redo-MitraClip was performed in both patients after partial detachment of the device from a flail posterior leaflet, at 4 and 5 months after the initial TEER procedure respectively. Of those undergoing MV surgery after TEER, one patient underwent MV replacement at 4 months after the index TEER procedure due to partial detachment of the device from a flail anterior leaflet, while MV repair was performed in another patient 4 months after MitraClip due to iatrogenic mitral stenosis and symptomatic heart failure.

**Table 3.**
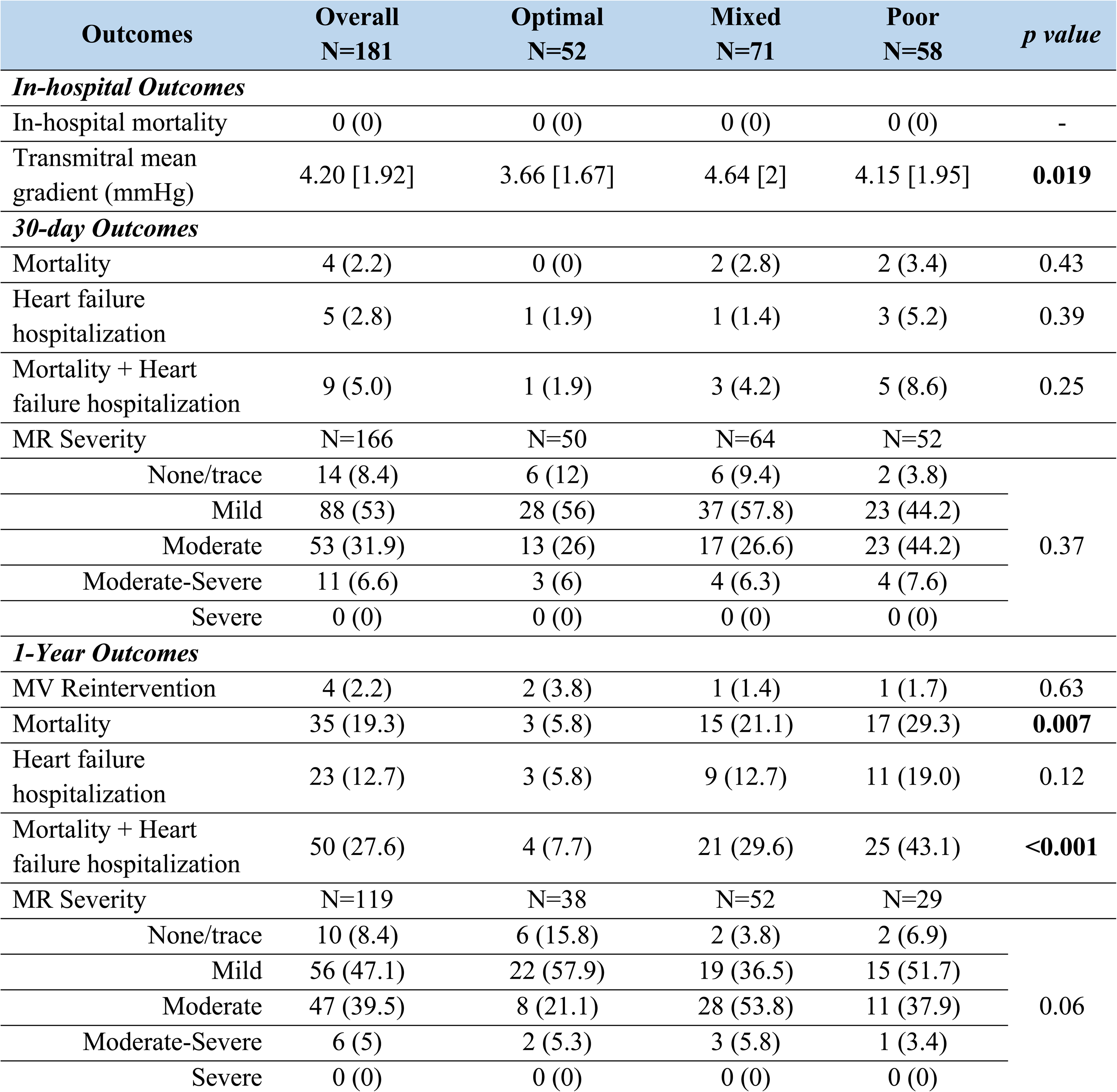
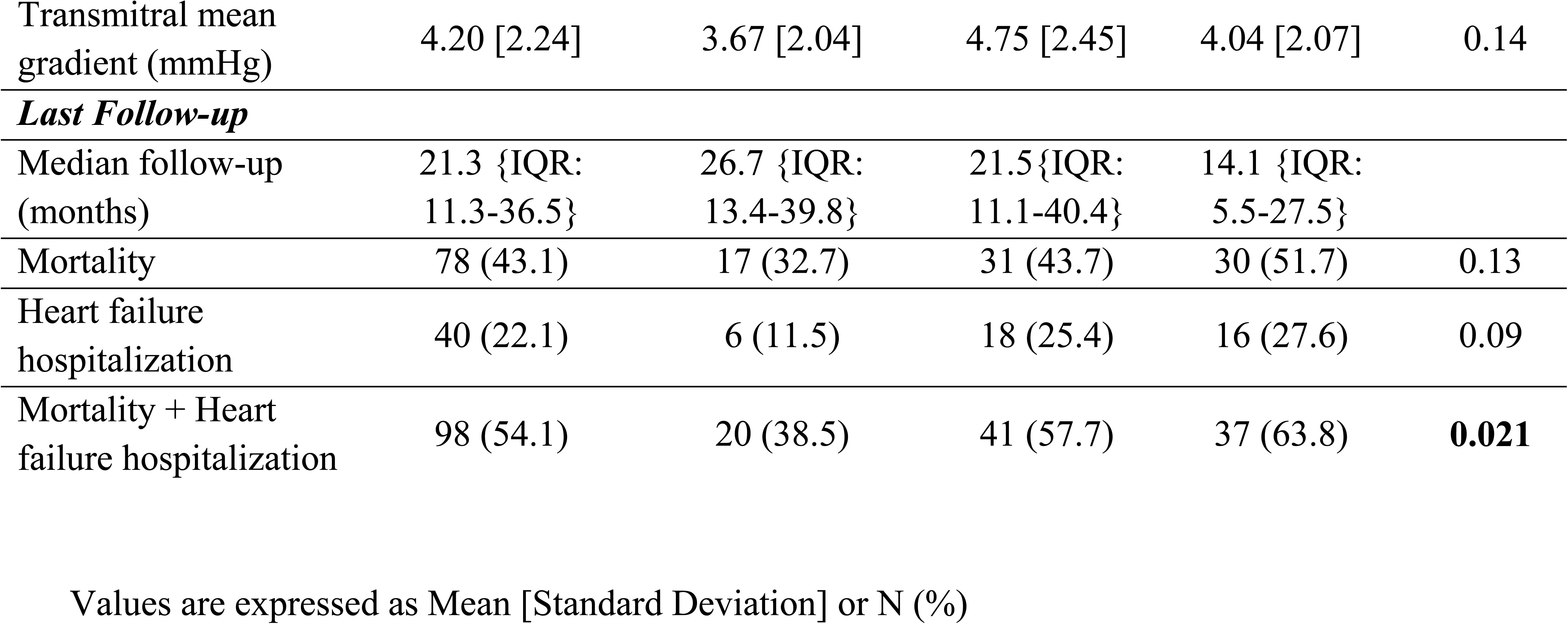
Outcomes.

The primary composite end-point of mortality and HFH occurred in 50 (27.6%) patients overall, with 1-year mortality and HFH rate of 19.3% and 12.7% respectively. On univariate analysis, 1-year mortality/HFH was associated with TR severity and lower LVEF at baseline, in addition to the severity of residual MR and v-wave after MitraClip implantation. (**Supplementary Table S2**). On multivariable cox regression analysis, residual mLAP [HR=1.073/mmHg (1.03-1.12)] and lesser degree of MR reduction [HR=0.65/grade (0.45-0.93)] and lesser S-VTI increment [HR=0.95/cm (0.91-0.99)] were independent predictors of 1-year mortality/HFH.

#### Intraprocedural Hemodynamic Profiling

For each independent predictor of the primary composite end-point, we identified the most predictive binary variable for the associated independent, continuous variable based on having the most robust area under the ROC curve in univariate modeling. We thus identified MR reduction by <3 grades (33.1%), S-VTI increment ≤8cm (33.9%), and residual mLAP >15mmHg (43.6%) as the most predictive thresholds that were independently associated with the composite of 1-year mortality/HFH. Based on the number of these predictors present, we created 3 intraprocedural hemodynamic profiles defined as Optimal (0 predictors), Mixed (1 predictor) and Poor (≥2predictors) profiles, which were seen in 28.7%, 39.2% and 32.0% of cases respectively **(Figure 2)**.

#### Optimal vs mixed vs poor hemodynamic profile

##### Baseline Characteristics

Compared to mixed/poor profile groups, the optimal profile patients were older (79.3 vs 77.1 vs 73.1 years, p=0.006) with higher baseline LVEF (55.7±13.2% vs 51.7±15.3% vs 46.6±14.4%, p=0.005), smaller LV dimension (left ventricular internal systolic diameter: 3.7±1.2 cm vs 3.7±1.2 cm vs 4.3±1.1 cm, p=0.009) and more severe MR at baseline (96.2% vs 88.7% vs 63.8%, p<0.001). There were no significant differences in other baseline clinical and echocardiographic characteristics between groups **(Table 1).** There were also no differences in the number and generation of MitraClip between groups **(Table 2).** In addition to color doppler findings, greater MR severity at baseline in the optimal profile was further evidenced by a higher proportion of PV systolic reversal (88.5% vs 62% mixed vs 31% poor, p<0.001), and a lower S-VTI (p<0.001) and peak Sv (p<0.001). However, there were no significant differences in baseline invasive mLAP and v-wave between groups. Comparison of pre-TEER, post-TEER and intraprocedural changes in PV Doppler and invasive hemodynamics among the 3 profiles is illustrated in **Figure 3 and Supplementary table S1.**

**Figure 3.**
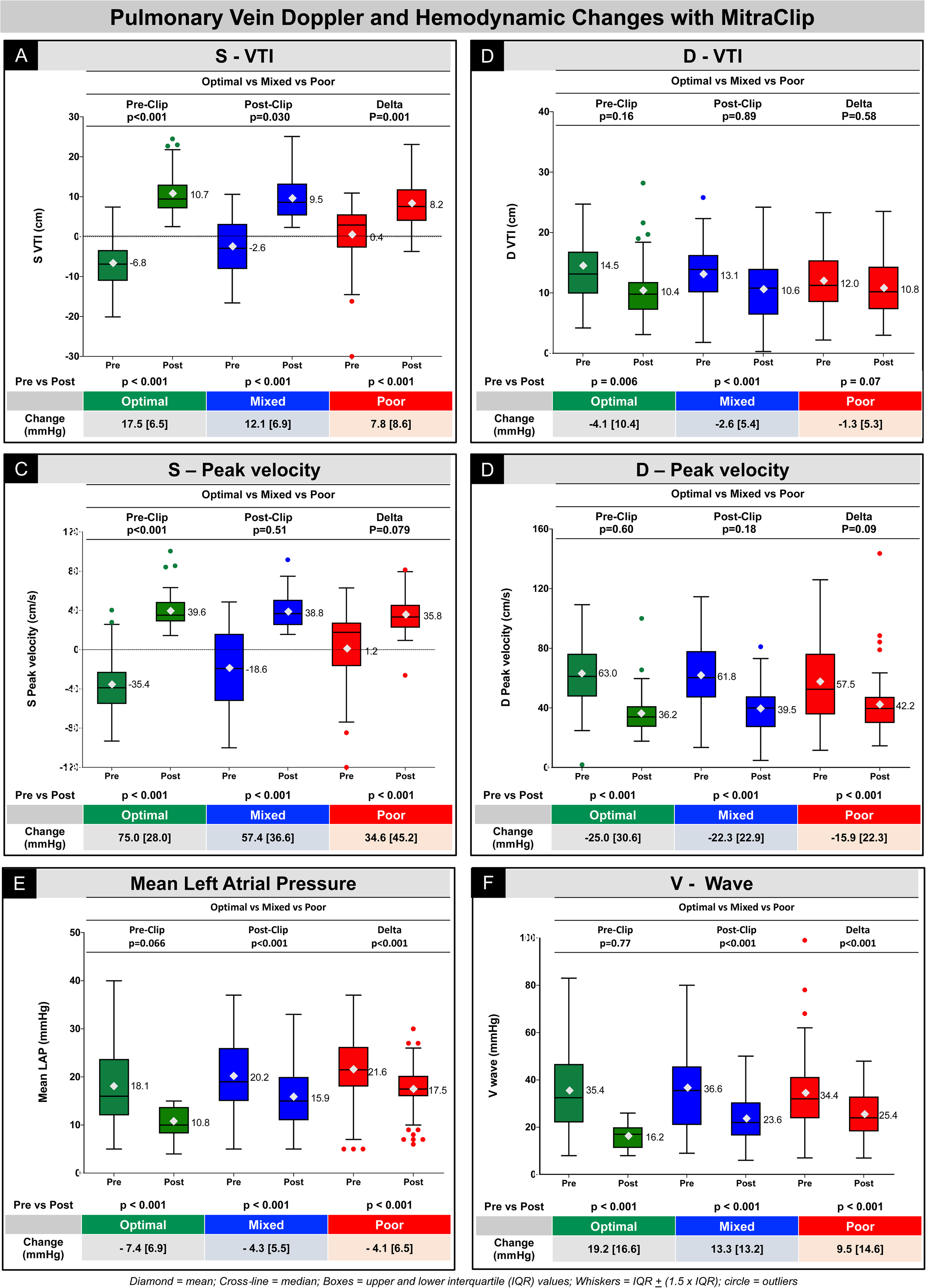
Pulmonary Vein Doppler and Hemodynamic Changes with MitraClip. Changes in Pulmonary Vein Doppler and Invasive Hemodynamics with MitraClip comparing velocity time integral of S-wave (A) and D-wave (B); peak velocity of S-wave (C) and D-wave (D) and Invasive mean left atrial pressure (E) and v-wave (F), between optimal versus mixed versus poor profiles.

##### Intraprocedural Changes with TEER

MitraClip implantation was associated with a significant reduction in MR severity accompanied by a significant change in both PV flow and invasive hemodynamics within each profile (all p<0.05). There was an increase in S-VTI and peak Sv, reduction in D-VTI and peak Dv, along with a significant reduction in invasive mLAP and v-wave (all p<0.05). However, the degree of MR reduction between groups did not reach statistical significance (3.2±0.4 vs 2.9±0.6 vs 2.2±0.8, p=0.21). When comparing the degree of change in each parameter after adjusting for baseline differences between groups, optimal profile had a greater increment in S-VTI (p=0.001), accompanied by a greater reduction in invasive mLAP (p<0.001) and v-wave (p<0.001) with mitral TEER.

##### Post-TEER Characteristics

≤Mild MR was seen in 79.5% of patients at the end of the procedure, with a significantly higher proportion in the optimal group (100% vs 83.1% vs 56.9%, p<0.001). While there were no differences in post-procedural PV wave morphology, peak Sv, peak Dv or D-VTI between groups, optimal profile had a higher post-procedural S-VTI (p=0.030), and lower invasive mLAP (p<0.001) and v-wave (p<0.001) after MitraClip.

#### Prognostic value of Intraprocedural Hemodynamic Profile

With a median follow-up of 21.3 months (IQR: 11.3-36.5) after mitral TEER, cumulative event-free survival was 72.0% at 1 year, and 60.1% at 2 years. Each component of the hemodynamic profile was associated with worse outcomes, with a lower 2-year cumulative event-free survival in patients with MR reduction <3 grades [52.8% vs 63.8%; HR 1.7 (1.1-2.6), p=0.011], S-VTI increment ≤8cm [43.1% vs 68.4%; HR 1.9 (1.2-2.8), p=0.004] and residual mLAP >15mmHg [43.7% vs 72.5%; HR 1.9 (1.3-2.8), p=0.002] **(Figure 4)**. 2-year cumulative event-free survival was higher in optimal profile (84.7% vs 55.5% mixed vs 43.3% poor, P<0.001), with an incremental risk of mortality/HFH with each profile overall [HR=1.75 per profile (1.34-2.29)], and within Primary [HR=1.64 per profile (1.15-2.36)] and Secondary MR [HR=1.77 per profile (1.17-2.68)] cohorts. Additionally, Hemodynamic profile was associated with overall survival alone, with a higher 2-year cumulative survival in optimal profile (89.0% vs 66.5% mixed vs 59.1% poor, P=0.003), and an incremental risk of mortality [HR=1.65 per profile (1.22-2.22)] with each profile.

**Figure 4.**
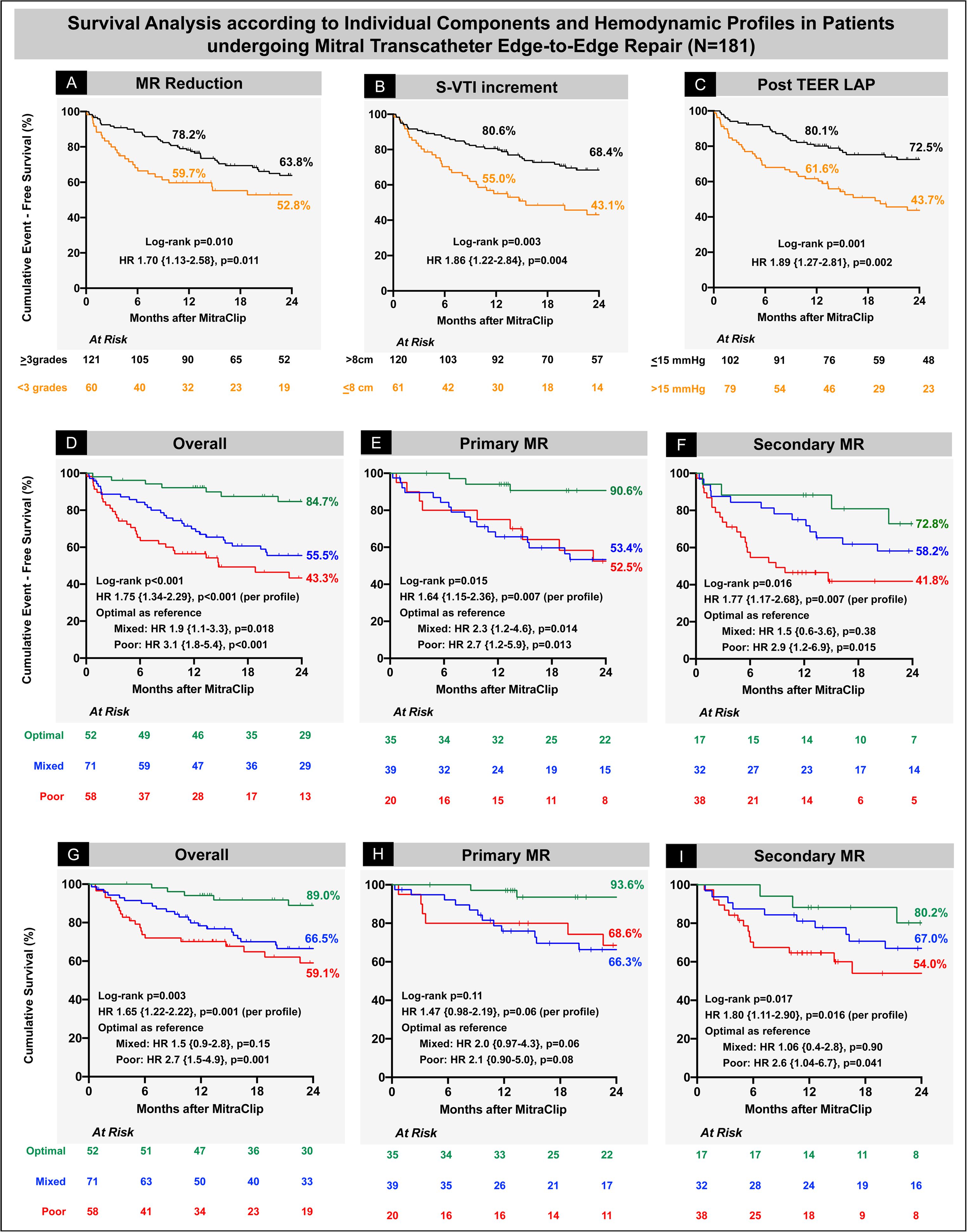
Survival Analysis according to Individual Components and Hemodynamic Profiles. Cumulative event-free survival was associated with each component of the hemodynamic profile, with lower 2-year cumulative event-free survival in patients with MR reduction <3 grades (A), S-VTI increment ≤8cm (B) and residual mLAP>15mmHg (C); and the intraprocedural hemodynamic profile with lower 2-year cumulative event-free survival overall (D), and within primary (E) and secondary MR (F) cohorts. Compared to patients with poor profile, optimal profile had superior survival overall (G), and within primary (H) and secondary MR (I) cohorts.

One-year mortality after mitral TEER was associated with prior stroke, mitral annular calcification severity, lower baseline LVEF and residual MR severity **(Figure 5)**. On multivariable Cox-regression, hemodynamic profile was an independent predictor of the 1-year composite of mortality and HFH [HR=1.98 per profile (1.21-3.25), p=0.007], in addition to baseline tricuspid regurgitation severity [HR=1.55 per grade (1.10-2.19), p=0.013], and post-procedural transmitral mean gradient >5mmHg [HR=2.32 per mmHg (1.17-4.61), p=0.016].

**Figure 5.**
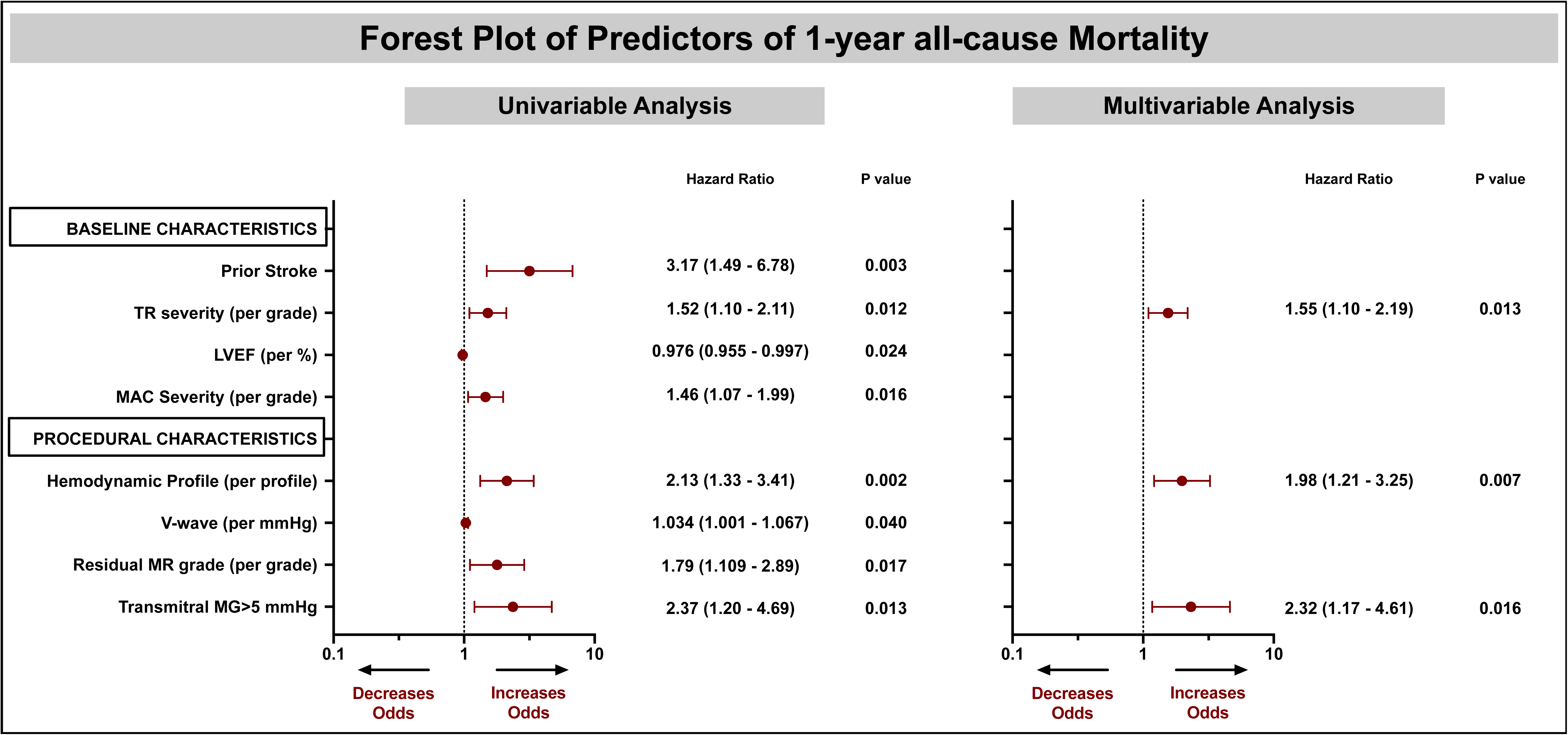
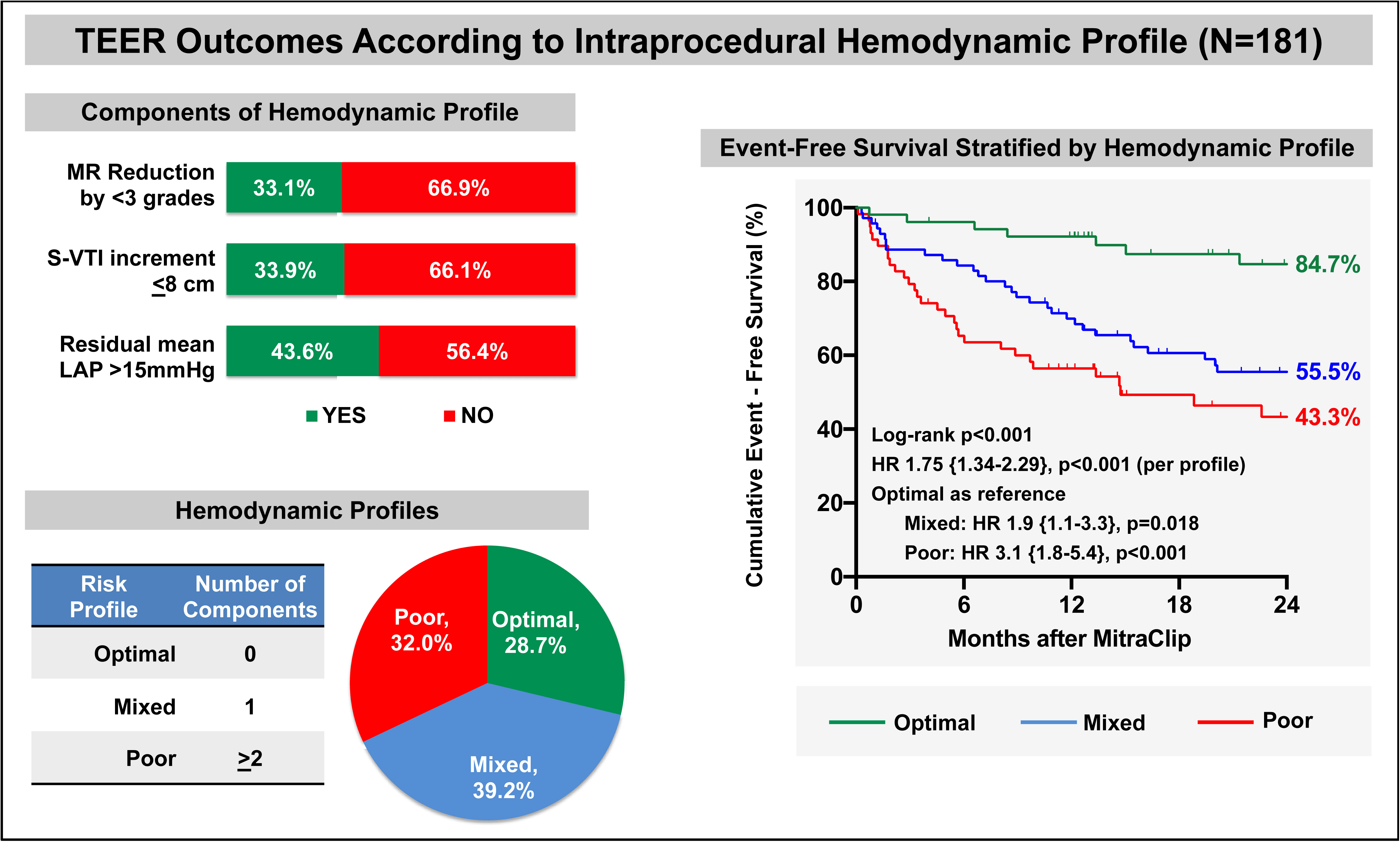
Predictors of Mortality after Mitral TEER. Forest plot showing univariable and multivariable predictors of 1-year mortality after Mitral TEER. **Central Illustration. TEER Outcomes According to Intraprocedural Hemodynamic Profile** Using optimal thresholds for change in pulmonary vein velocity-time-integral of S-wave (S-VTI increment ≤8cm), MR reduction (<3grades), and residual mean left atrial pressure (>15mmHg), three hemodynamic profiles were developed: optimal, mixed, or poor with 0, 1 or ≥2 components respectively. Compared to mixed / poor groups, optimal profile was associated with higher 2-year cumulative event-free survival (84.7% vs 55.5% vs 43.3%, log-rank p<0.001), with an incremental risk of mortality and heart failure hospitalization [HR: 1.75 per profile (1.34-2.29)] with each profile. Hemodynamic profile was an independent predictor of 1-year mortality after TEER [HR=1.98 per profile (1.21-3.25), p=0.007].

## Discussion

In this retrospective single center study consisting of ∼50% primary MR patients, the clinical applicability of intraprocedural hemodynamic profile was examined which for the first-time integrated real-time changes in non-invasive doppler and invasive hemodynamics to study outcomes after MitraClip. Our key findings are as follows: First, cumulative event-free survival was 72.0% at 1-year after TEER; Residual mLAP in addition to a lesser degree of MR reduction and lesser S-VTI increment were independent predictors of the primary composite end-point of 1-year mortality and HFH. Second, MR reduction <3grades, S-VTI increment ≤8cm, and residual mLAP >15mmHg were the most predictive thresholds of the primary composite endpoint. Third, intraprocedural hemodynamic profiles conceived based on the number of predictors present showed that optimal (0 predictors), mixed (1 predictors) and poor (≥2 predictors) profiles were observed in 29%, 39% and 32% of patients respectively. Fourth, optimal intraprocedural hemodynamic profile was associated with superior outcomes overall, and within primary and secondary MR cohorts. There was an incremental risk of both mortality and mortality/HFH with each profile. Finally, intraprocedural hemodynamic profile was an independent predictor of 1-year mortality after Mitral TEER, in addition to baseline TR severity and post-procedural transmitral mean gradient >5 mmHg.

### Prognostic value of PV Flow in mitral TEER

PV profile is a valuable tool in the assessment of MR severity and its hemodynamic impact. PV flow has systolic (S) and diastolic (D) waves, with the former being influenced by changes in LAP and LA relaxation. MR severity is associated with a progressive reduction in the peak S velocity, with some patients having the characteristic systolic reversal which has high specificity for severe MR.^16^ While many operators believe that a change in PV flow from systolic reversal at baseline to systolic predominance after MitraClip implantation is an indicator of procedural success, this has not been systematically studied. There are also limited data on the prognostic value of changes in intraprocedural PV flow during Mitral TEER.

One of the earliest studies that examined changes in PV flow during MitraClip showed an immediate increase in peak Sv and Sv/Dv ratio in response to MR reduction. ^17^ Lower post-procedural S-VTI/D-VTI ratio (≤0.72) has been associated with more significant residual MR and higher incidence of MACE at 12 months.^11^ Improvement in PV morphology after mitral TEER in a more recent study was associated with superior long-term survival and freedom from rehospitalization.^12^ However, it is unclear what degree of improvement in the S wave defines procedural success and portends superior outcomes. This is particularly important in patients with secondary MR undergoing mitral TEER where systolic blunting may be present at baseline. Additionally, S-VTI may be a more reliable and reproducible measure of procedural success since it is not affected by changes in vessel diameter or heart rate. In our study, a lower increment in S-VTI was an independent predictor of 1-year mortality and HFH after TEER, with S-VTI increment of ≤8cm being the most predictive threshold associated with worse outcomes.

### Intraprocedural Hemodynamic Profiling in Mitral TEER

The primary objective of TEER is to reduce MR to ≤ mild without causing significant mitral stenosis. While this remains the Achilles heel of the TEER device technology, operators have adopted adjunctive tools to assess immediate hemodynamic response to TEER such as continuous invasive LAP monitoring made possible with the newer MitraClip G4 system, and changes in PV flow. However, there are conflicting data on the prognostic value of residual mLAP and degree of MR reduction. For example, reduction in LAP was an independent predictor of all-cause mortality and HFH after TEER in an observational study ^18^, while another study showed that final LAP was not associated with adverse outcomes.^12^ Similarly, contrary to prior data that showed a significant association between >moderate residual MR and 1-year mortality ^19, 20^, Corrigan et al reported that residual MR severity did not predict worse outcomes after TEER.^12^ In our study, lesser MR reduction and higher residual mLAP were independent predictors of 1-year mortality and HFH after TEER, with MR reduction <3 grades and residual mLAP >15mmHg being the most predictive threshold associated with worse outcomes.

Furthermore, hemodynamic and echocardiographic studies exploring outcomes in mitral TEER have been performed in isolation, and no prior studies have integrated non-invasive doppler and invasive hemodynamic changes to study long-term outcomes after MitraClip. A recent study by Sato et al that incorporated invasive residual mLAP following mitral TEER and residual MR on postprocedural day-1, reported-on outcomes based on 3 hemodynamic profiles and showed that patients with an optimal hemodynamic profile that consisted of residual mLAP ≤15 mmHg and residual MR ≤1 had the highest 1-year survival rate.^10^ While the concept of hemodynamic profiling was intriguing, the profile itself was applicable only at the time of discharge and not during the TEER procedure. The study also predominantly included primary MR patients (83%), and changes in PV flow were not evaluated.

Hemodynamic profiling by integrating non-invasive doppler and invasive hemodynamic changes during the MitraClip procedure may provide real-time intraprocedural decision guidance to further optimize long-term clinical outcomes. In our study, MitraClip therapy was associated with a significant reduction in MR severity (97.8% <moderate MR post-clip), increase in S-VTI (from -2.8±7.4 to 9.4±5.1 cm, p<0.001) and reduction in invasive mLAP (from 20±7.8 to 14.9±5.7 mmHg, p<0.001), all of which were independently associated with superior outcomes. We used the most predictive thresholds for each predictor to develop 3 hemodynamic profiles based on the number of predictors present - optimal (0 predictors), mixed (1 predictors) and poor (≥2 predictors). We found that there was an incremental risk of both mortality and mortality/HFH with each profile. Additionally, intraprocedural hemodynamic profile was an independent predictor of 1-year mortality after TEER.

Despite being older, optimal profile patients in our study had superior outcomes compared to the mixed/poor groups. The optimal group had higher baseline LVEF, smaller LV dimension and more severe MR at baseline as corroborated by a higher proportion of PV systolic reversal and lower S-VTI and peak Sv. This group likely represents patients with the least LV remodeling, and those deriving the greatest benefit from MR reduction therapy. This is further evidenced by a significantly greater reduction in mLAP and v-wave in the optimal group, despite no differences in baseline mLAP between groups. Additionally, our study cohort had nearly even distribution between primary and secondary MR etiologies, and optimal profile patients had superior outcomes within each etiology which further strengthens the discrimination of the profile. These findings are particularly noteworthy in the light of recent studies that highlight the need to identify responders to mitral TEER therapy.^21^ Operators often face situations where MR reduction is suboptimal and optimization prior to release is critical particularly in patients with challenging anatomies, and intraprocedural hemodynamic profiling may be helpful in providing real-time decision guidance.

### Predictors of one year mortality

In our study cohort consisting of ∼50% primary MR patients, pre-procedural TR severity and TMPG >5 mmHg at the time of discharge were independent predictors of 1-year mortality after TEER, in addition to intraprocedural hemodynamic profile. Our findings are consistent with reports from the COAPT study and a recent meta-analysis where patients with >moderate TR at baseline undergoing TEER had worse outcomes.^5, 22^ However, prognostic impact of TMPG on outcomes after TEER has been a matter of debate. Koell et al found that pre-discharge TMPG ≥ 5 mmHg was an independent predictor of death or HFH in those with primary MR (37%) but not secondary MR (63%).^23^ In a post hoc analysis form the COAPT trial, higher TMPG did not adversely affect the 2-year outcomes of all-cause mortality or HFH.^24^ Our findings underscore the importance of integrating real-time changes in PV flow, invasive mLAP, MR severity and TMPG in addition to baseline TR severity which could be of paramount help in guiding decisions for future mitral TEER procedures.

## Study limitations

Our study has certain limitations. First, the retrospective nature of this study at a single institution has inherent limitations and biases, including time bias as different TEER device generations were included. Second, we excluded 37% of patients who underwent mitral TEER during the study period due to lack of high-quality PV Doppler tracings; only those with analyzable PV Doppler tracings both before and after MitraClip deployment were included. While this may have introduced a selection bias, outcomes in those excluded (1-year mortality 23.8%; 1-year mortality/HFH 28.6%) were similar to the primary study cohort. Third, MR may affect the PV flow differentially in the left vs right PV especially in patients with eccentric jets. However, we interrogated the same PV before and after MitraClip implantation in each patient and procedural success was based on change in flow in the same PV. Fourth, >50% of our patients had a history of atrial fibrillation that likely reduces systolic PV flow regardless of MR severity due to lack of atrial contraction and relaxation. However, there were no differences in prior atrial fibrillation across profiles (p=0.78) and we focused on the acute change in PV Doppler peri-procedurally. Finally, there was no independent echocardiographic core laboratory to assess the echocardiographic parameters before and after the procedure and a single experienced echocardiographer (PW) performed all echocardiographic evaluation.

## Conclusion

Intraprocedural hemodynamic profile is a novel prognostic stratification tool for patients undergoing TEER that integrates real-time invasive hemodynamic and non-invasive Doppler changes during the MitraClip procedure. Hemodynamic profiling using changes in TEE-derived PV flow, MR severity and invasive LAP may provide real-time intraprocedural decision guidance to optimize long-term clinical response in our pursuit of surgery-like outcomes with TEER. Further longer-term studies are warranted to validate our findings in a larger independent cohort.

### Perspectives

#### What is New?

- Impact of various hemodynamic and echocardiographic changes on outcomes after mitral TEER has been studied in isolation; No prior studies have integrated real-time changes in non-invasive Doppler and invasive hemodynamics to study long term outcomes after MitraClip.
- Intraprocedural Hemodynamic profiling based on the degree of MR reduction, improvement in TEE-derived PV forward flow and invasive residual mLAP predicts mortality and composite of mortality/HFH after mitral TEER.

#### What are the clinical Implications?

- Further in-depth studies integrating real-time changes in non-invasive doppler and invasive hemodynamic are necessary to refine prognostic stratification and provide intraprocedural decision guidance during mitral TEER.
- Longer-term studies in both Primary and Secondary MR patients are warranted to validate our findings in a larger independent cohort.

## Data Availability

The data that support the findings of this study are available on request from the corresponding author [SG].

## Abbreviations

LAP: Left atrial pressure
MR: Mitral Regurgitation
MV: Mitral Valve
PV: Pulmonary Vein
TEE: Transesophageal Echocardiography
TEER: Transcatheter edge-to-edge repair
VTI: Velocity time integral

---

**Supplementary Table S1.**
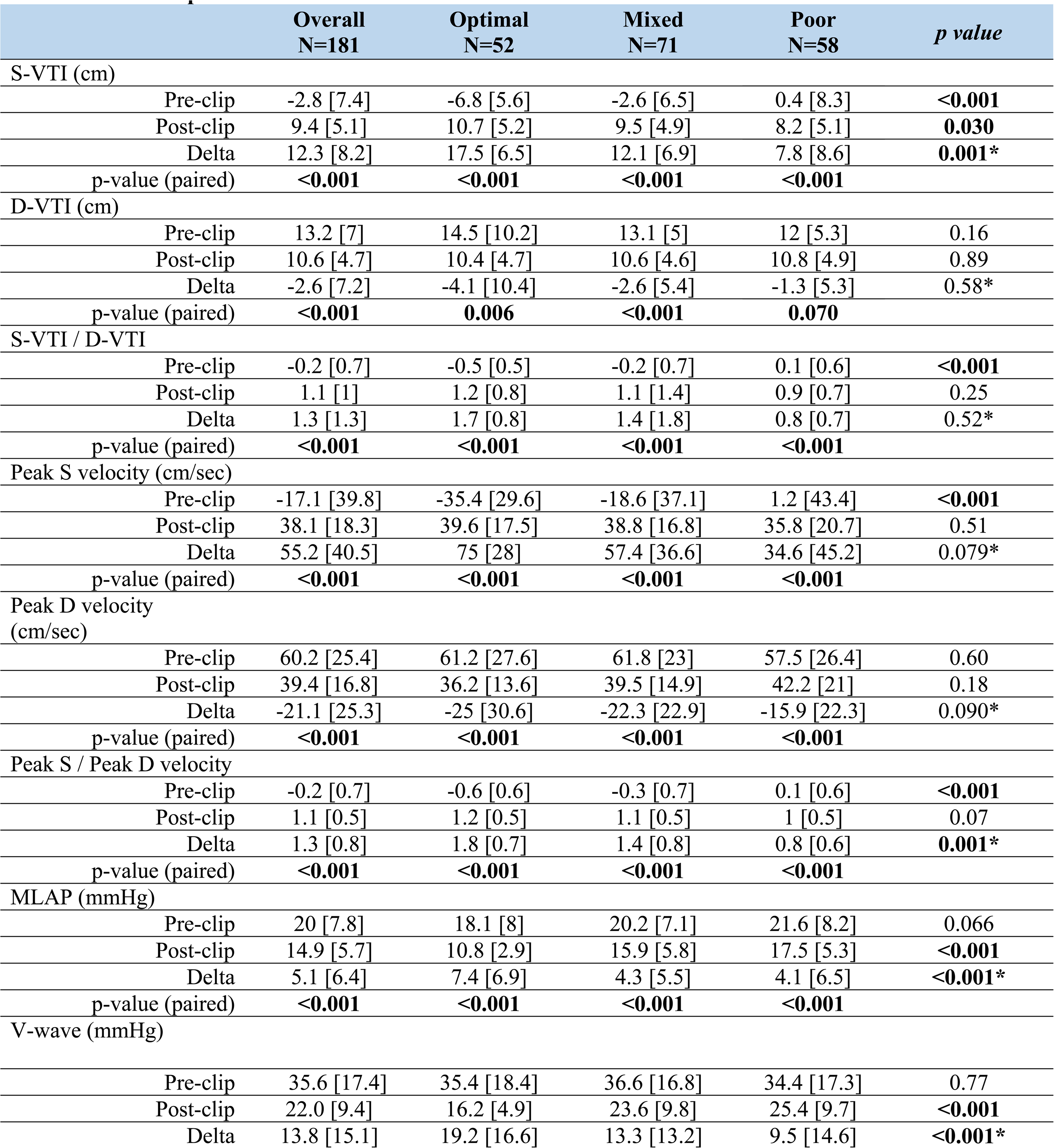
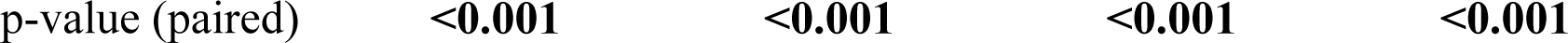
Pulmonary Vein Doppler and Hemodynamic Changes with MitraClip.

**Supplementary Table S2.** Intraprocedural Predictors of the primary composite endpoint of 1-year Mortality and Heart Failure Hospitalization after MitraClip.

|  | Univariable Analysis |  |  |  | Multivariable Analysis |  |  |  |
| --- | --- | --- | --- | --- | --- | --- | --- | --- |
|  | Hazard Ratio | 95% Confidence interval |  | p-value | Hazard Ratio | 95% Confidence interval |  | p-value |
|  |  | Lower | Upper |  |  | Lower | Upper |  |
| Pre-TEER |  |  |  |  |  |  |  |  |
| TR severity (per grade) | 1.428 | 1.093 | 1.866 | 0.009 |  |  |  |  |
| LVEF (per %) | 0.968 | 0.950 | 0.985 | <0.001 |  |  |  |  |
| Change with TEER |  |  |  |  |  |  |  |  |
| Delta S-VTI (per cm) | 0.942 | 0.906 | 0.980 | 0.003 | 0.951 | 0.913 | 0.990 | 0.015 |
| Delta peak Sv (per m/sec) | 0.990 | 0.982 | 0.998 | 0.010 |  |  |  |  |
| Delta MR (per grade) | 1.703 | 1.192 | 2.435 | 0.003 | 0.65 | 0.45 | 0.93 | 0.019 |
| Post-TEER |  |  |  |  |  |  |  |  |
| Mean LAP (per mmHg) | 1.074 | 1.028 | 1.122 | 0.001 | 1.073 | 1.028 | 1.120 | 0.001 |
| V-wave (per mmHg) | 1.042 | 1.015 | 1.069 | 0.002 |  |  |  |  |
| MR severity (per grade) | 1.615 | 1.079 | 2.417 | 0.020 |  |  |  |  |

**Supplementary Figure S1:**
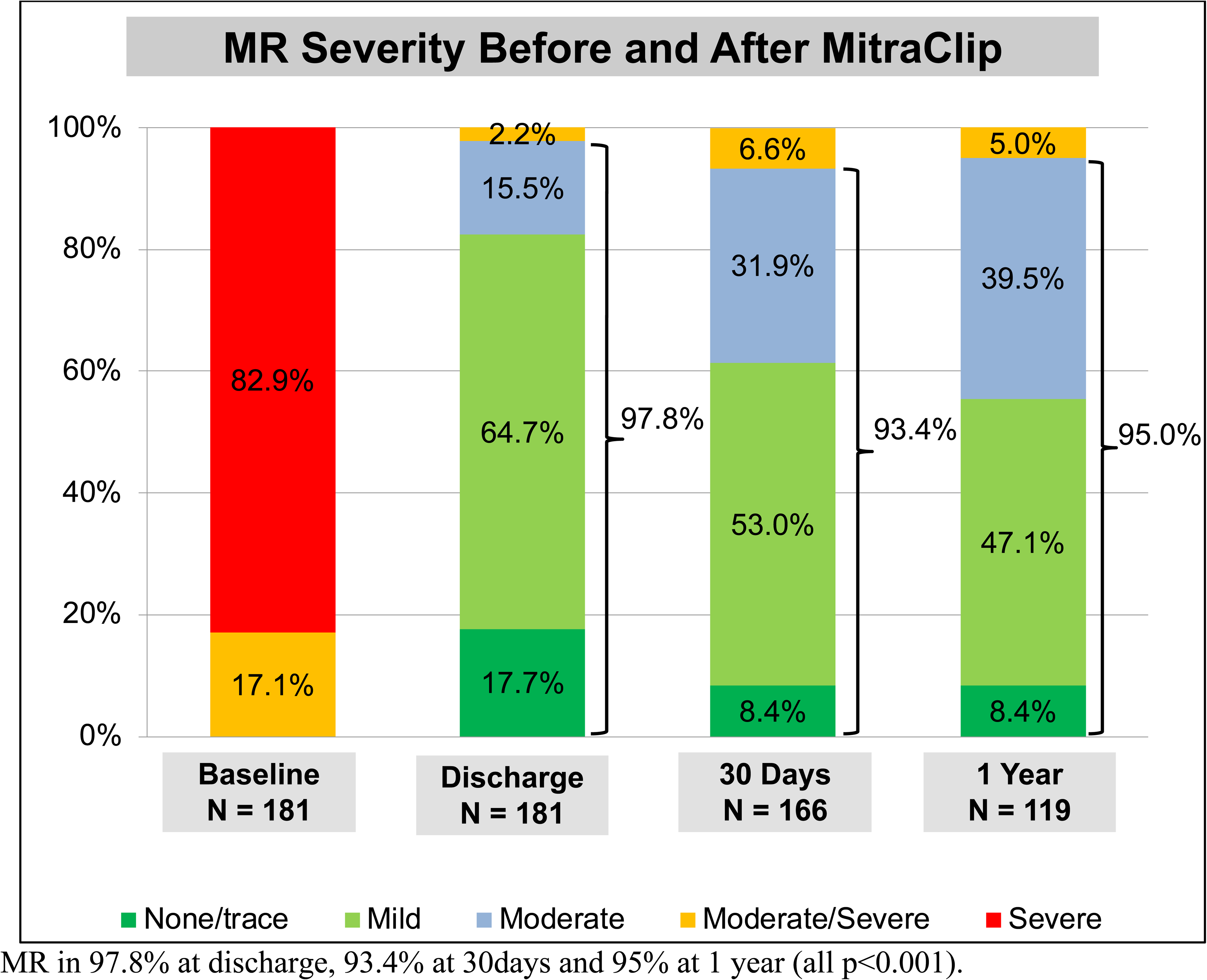
Mitral Regurgitation Severity Before and After MitraClip. There was a significant reduction in MR severity after MitraClip implantation with ≤moderate MR in 97.8% at discharge, 93.4% at 30days and 95% at 1 year (all p<0.001).

